# Analyses of biomarkers for tremor using local field potentials recorded from deep brain stimulation electrodes in the thalamus

**DOI:** 10.1101/2025.01.03.24319798

**Authors:** Karthik Kumaravelu, Stephen L. Schmidt, Yi Zhao, Allison Vittert, Brandon D. Swan, Chintan S. Oza, Jennifer J. Peters, Kyle T. Mitchell, Dennis A. Turner, Warren M. Grill

## Abstract

**Background:** Deep brain stimulation (DBS) of the ventral intermediate nucleus (VIM) of the thalamus (TH) is an effective therapy for suppressing tremor. One of the critical challenges to optimizing VIM-DBS therapy is the lack of robust neural biomarkers that correlate well with tremor.

**Objective:** We conducted intraoperative local field potential (LFP) recordings from DBS electrodes placed in the TH (including VIM and ventralis oralis posterior-VOP) to quantify biomarkers of tremor. We used computational modeling to understand the biophysical basis of the recorded LFP signal.

**Methods:** We simultaneously recorded intraoperative TH LFP and tremor from the hand dorsum (34 participants) and during DBS at different frequencies (16 participants). Then, we simulated the effects of DBS and spatial distribution of tremor cells on calculated LFPs in a TH model.

**Results:** There was a moderate correlation between tremor and LFP spectral power in the theta and alpha bands (r = 0.445 and 0.389, respectively). There was a strong correlation between tremor and peak coherence between LFP and tremor signal (r = 0.559). Postural tremor was decoded from the LFP signal with an area under curve of ∼0.7. High frequency DBS reduced spectral power in the theta and alpha bands and tremor could be decoded from the LFP spectral power in the presence of DBS (0.429 goodness of fit R^2^). The theta power in the simulated LFP signal varied substantially with the specific location of the bipolar contact pair of the DBS electrode used for the LFP recordings as well as the spatial distribution of tremor cells.

**Conclusions:** Theta power alone was not sufficient for prediction of tremor control. Simulations indicated that the number and distribution of tremor cells surrounding the DBS lead may explain the lack of strong correlation between tremor and theta power.

*HIGHLIGHTS:*

- Recorded intraoperative thalamic LFP in patients with ET, PD, or multiple sclerosis.
- Moderate correlation between tremor and spectral power in theta/alpha bands.
- Strong correlation between tremor and LFP-Accelerometer coherence.
- Tremor can be decoded from VIM-LFP with an AUC of 0.7.
- Thalamic model indicated theta power dependent on tremor cell distribution.

## INTRODUCTION

Deep brain stimulation (DBS) of the ventral intermediate nucleus (VIM) of the thalamus (TH) is a surgical therapy effective in managing tremor in essential tremor (ET), multiple sclerosis, and tremor-dominant Parkinson’s disease (PD) [1–3]. Despite the widespread use of VIM-DBS, its mechanism of action remains unclear [4]. Further, current DBS systems to treat tremor are open-loop, i.e., stimulation is continuous and invariant [5, 6]. One of the key challenges to developing “closed-loop” or self-adjusting DBS for managing tremor is identifying physiological, internal neural biomarkers that correlate with tremor control. Although there are successful demonstrations of closed-loop control of DBS for tremor using external sensors [7–9] or cortical sensing [10–13], it is highly desirable to achieve closed-loop control with the same lead used for stimulation to avoid the necessity of additional implanted electrodes or the use of external hardware.

Microelectrode recordings show cells oscillating in the theta band (tremor cells) distributed across the TH nuclei (VIM + ventralis oralis posterior – VOP + ventral caudal – VC) [14–16], and macroelectrode recordings in these nuclei enable detection of theta band local field potentials (LFP) in small case series (7 to 14 participants) [17–20]. Tremor cells have been implicated in the pathogenesis of tremor, but it is not clear whether low-frequency oscillatory activity in LFPs correlates well with tremor magnitude across a large cohort of patients nor how these signals are influenced by DBS [3]. Coherence between LFPs and tremor (via electromyography – EMG) was found to lag the onset of tremor in patients with ET [21]. However, the correlation between coherence and tremor magnitude has not been shown systematically across many patients. Although a study has demonstrated closed-loop control of DBS using VIM LFPs [22], no prior study has quantified the modulation of VIM LFP spectral power and tremor at different DBS frequencies.

To address these important gaps, we recorded LFPs from the TH (including VIM and VOP) using a bipolar pair of a four contact DBS lead in ET, multiple sclerosis, and PD patients undergoing surgery for either lead implantation or pulse generator replacement (34 participants). We simultaneously recorded tremor using an accelerometer placed on the hand dorsum. There was a moderate correlation between tremor and low-frequency LFP spectral power in the alpha and theta bands. However, there was a strong correlation between tremor and peak coherence between LFPs and acceleration. We decoded postural tremor from the LFP signal with an area under curve (AUC) of ∼0.7. High frequency DBS (∼130 Hz) reduced the low frequency spectral power compared to baseline and postural tremor was decoded from LFP signals recorded in the presence of DBS. Subsequently, we used computational modeling to understand the biophysical basis of the recorded LFP signals. The low-frequency spectral power of the simulated LFP varied considerably with parameters including electrode location, the spatial distribution of tremor cells across the TH, and the proportion of tremor cells relative to non-tremor cells. These sensitivities might account for the differences in LFP spectral power across participants due to inherent variability in surgical implantation/accuracy and the only moderate correlation with tremor.

## METHODS

### Human participants and intraoperative experiments

All procedures were approved by the Institutional Review Boards of Duke University Health System and Emory University Hospital, and all participants provided written informed consent. 34 patients undergoing surgery for thalamic DBS implant or implanted pulse generator replacement participated in the study (Fig. 1A and Participant Data Table in Supplementary Materials). Of the 34 participants, accelerometry data from 13 participants was previously analyzed and presented in [23]; however, unique analyses are presented here. Participants withdrew from anti-tremor medications for at least 12 hours prior to surgery. Methodological details are available for pulse generator replacement subjects in [24] and for implant subjects in [23]. Briefly, leads were externalized and connected to a custom amplifier and DBS platform [23, 25]. Voltage-controlled monopolar DBS was delivered using Medtronic 3387 or 3389 leads (Medtronic PLC, Dublin, Ireland). The attending neurologist determined the stimulation amplitude and pulse width sufficient for therapeutic benefit. The pulse width was typically held at either 60 or 90 μs, and the stimulation amplitude was varied to determine the therapeutic window. This clinically derived, safe stimulation amplitude was then used for subsequent LFP recordings. LFPs were recorded using differential sensing around the stimulation contact (Participant Data Table in Supplementary Materials). The LFP was amplified by two or three SR560 amplifiers connected in series (Stanford Research Systems, Sunnyvale, CA) and digitized by a NIDAQ (National Instruments, Austin, TX). Acceleration was collected with an CXL04LP3 accelerometer (Crossbow Technology, Milpitas, CA) and digitized by the NIDAQ (Fig. 1B). Tremor assessment occurred for 20 s beginning after 30s of DBS (Fig. 1C). If tremor was minimal, arm maneuvers and other posture changes were performed to elicit tremor. In a smaller cohort n = 16, DBS frequencies of 2, 5, 10, 50, and 130 Hz were applied for 60 s with DBS off preceding each experimental DBS frequency.

**Figure 1:**
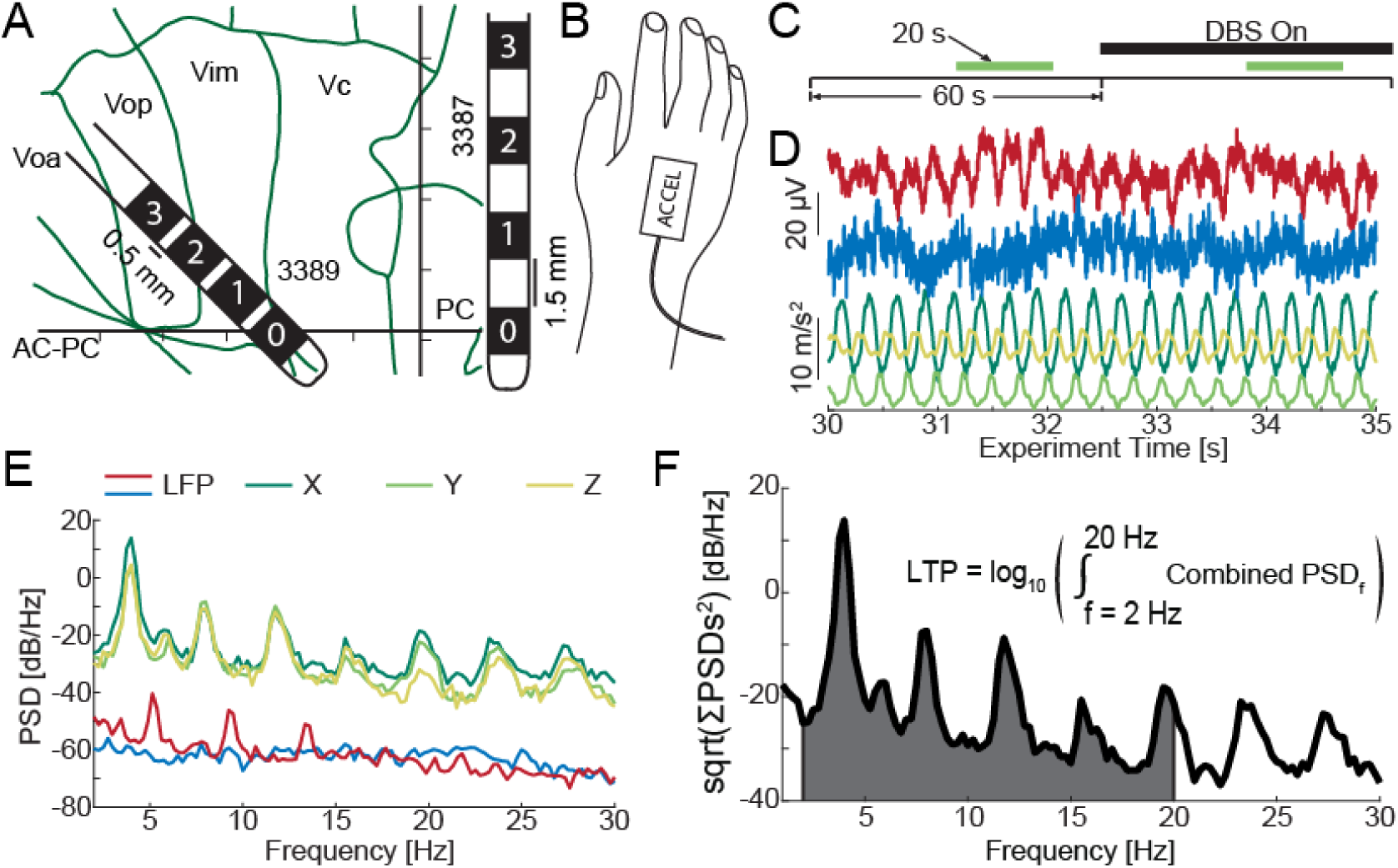
Intraoperative recording of local field potentials (LFP) and tremor in participants undergoing surgery for thalamic DBS. A. Map of thalamus (TH) based on (Brodkey et al. 2004). Tick marks indicate 2 mm. A typical placement of a Medtronic 3389 lead is shown. A scale representation of a 3387 lead is shown to the right. B. A tri-axial accelerometer was placed on the back of the hand to quantify tremor. C. Timing of a pair of trials. In the first 60 s LFP and acceleration were recorded without deep brain stimulation (DBS), and in the second 60 s LFP and acceleration were recorded during DBS. After 30 s tremor was assessed for 20 s (green) D. (Red) LFP from the participant with the largest fraction of LFP in the theta band. Simultaneous LFP (blue) and acceleration traces (green through yellow) during a 5 s epoch of a tremor measurement period of a typical participant with high tremor. E. The power spectral densities of each axis of acceleration of the typical participant and simultaneous LFP (blue). (Red) LFP PSD from participant with the largest fraction of LFP in the theta band. F. The combined power spectral density from the acceleration spectra in E. This spectrum was then integrated from 2 to 20 Hz and log transformed to yield log tremor power (LTP).

We localized DBS electrodes using BrainLab software (Brainlab AG, Munich Germany) by merging of the pre-op MRI and the postop CT. The anterior commissure (AC), posterior commissure (PC), and midline were identified, and the tip of the lead mapped to AC-PC coordinates. Sensing and stimulation contact locations were calculated by backtracking along the electrode trajectory given known contact spacing. We mapped these specific coordinates to a digitized Schaltenbrand-Wahren atlas [26]. To standardize measurements, we determined the length of the AC-PC line for each patient and scaled the anterior distances of contacts to correspond to a standard AC-PC length of 24 mm. Details about the DBS lead type and location of contact 0 with respect to the anatomical landmarks across the 34 participants are provided in the Participant Data Table in Supplementary Materials. The DBS electrode placement was intended that the most distal contacts (contacts 0 and 1) were targeted towards the posterior base of the VIM, with the proximal contacts (contacts 2 and 3) located in the VOP (Fig. S1A). The lateral distance depended on the width of the third ventricle (typically 10-12 mm). The mean location of the positive contact used for recording LFP across participants was 0.63 mm inferior to AC-PC line and 4.88 mm anterior to the PC point (Fig. S1B). Similarly, the mean location of the negative contact was 2.62 mm above the AC-PC line and 7.02 mm anterior to the PC point (Fig. S1B).

### Analysis of LFP and acceleration signals

LFP and acceleration data were analyzed using custom Matlab scripts (Mathworks, Natick, MA; Fig. 1D). In the stimulation cohort, DBS artifacts were removed from the LFP by first removing individual pulses by linear interpolation between 0.2 ms before to 1.5 ms after each pulse. Template subtraction was then applied using the 20 200 ms templates closest in time to the current 200 ms epoch. Template-subtracted LFPs were lowpass filtered (Butterworth 4^th^ order, noncausal) and down-sampled to 500 Hz to match the sampling rate of the accelerometer. When comparing between DBS off and DBS on, trials of DBS off were blanked and template subtracted as though the stimulation pattern of the paired DBS on trial had been applied. Power spectral densities were calculated using Welch’s algorithm during the 20 s motor assessment window (Fig. 1E). Tremor power was calculated from the power spectral density of each accelerometry axis summed across dimensions using the Pythagorean theorem and integrated from 2-20 Hz to yield a single value (Fig. 1F). Principal component analysis was used to reduce the tremor signal to a single dimensional time series. Magnitude squared coherence (MS coherence) was calculated during the 20 s motor assessment window between z-scored LFP and the first principal component of the tremor data. The window size was 2048 samples with 50% overlap resulting in a significance threshold of ∼0.146. Coherence was calculated for each frequency from 1-30 Hz with 0.25 Hz resolution. Those LFP-accelerometry pairs that exceeded the threshold were then compared to the 95^th^ percentile of 200 time-shifted shuffled controls. Frequencies exceeding both the standard significance criterion and the shuffle control were deemed to exhibit significant coherence. The peak tremor frequency was determined as the frequency with the largest power in the 2-7 Hz band. The tremor harmonic band was calculated as twice the tremor frequency ± 2 Hz. Power in the delta (1-4 Hz), theta (4-8 Hz), alpha (8-12 Hz), beta (13-30 Hz) and tremor harmonic bands was calculated from the LFP. Normalized power in each frequency band was normalized by the total power between 1-30 Hz. Due to a 24 Hz artifact observed in some of the data, we did not include 23.5-24.5 Hz in the beta and broadband power calculations.

Correlations are reported using Spearman R and p values. Multiple linear regression (MLR) was conducted on all power data using the Matlab function regress(). Reported R^2^ and p values were taken directly from the ‘stats’ vector returned by regress(). Trials with high tremor were classified by generalized linear models (GLM) for binomial distributions trained using the Matlab function fitglm(). Receiver operating characteristic (ROC) curves and AUC were calculated using the matlab function perfcurve(). Statistical significance of changes in tremor and LFP power bands by DBS were analyzed by JMP Pro (SAS Institute, Cary, NC) with the Full Factorial Repeated Measures Analysis of Variance (RMANOVA) add-in. The effect of each frequency of DBS on each power band was calculated using (RMANOVA) with Participant ID as a random factor. Two-sided RMANOVAs were used with one exception. Change in log tremor power (LTP) by DBS in low LTP participants was one-sided. The *a priori* hypothesis was that DBS of less than 100 Hz would increase LTP and DBS of greater than 100 Hz would decrease LTP. Power data were entered after a log_10_ transformation; therefore the resulting RMANOVAs assume a log-normal distribution of the residuals.

### Computational model of thalamus

The computational model of TH DBS comprised three regions: (1) VC nucleus, (2) VIM and (3) VOP, each represented as a rectangular prism with dimensions 3.5 mm x 4.5 mm x 8 mm (Fig. S2A). The models of individual neurons used in this study are described in detail elsewhere [27]. Briefly, the model comprised a multicompartment thalamocortical relay (TCR) neuron with somatic, dendritic, and axonal compartments (Fig. S3). The model comprised a total of 5000 TCR neurons with the soma of each TCR neuron constrained within the rectangular prism (Fig. S2A). The TCR neuron received excitatory inputs from the cortex (CTX) and the cerebellum (CER) represented using virtual axons, i.e., the synapses on the dendrites of the TCR neuron were directly driven with pulsatile inputs (Fig. S2B). The CER inputs comprised a burst of pulses at a rate of 5.8 Hz [11 pulses at an interpulse interval (IPI) of 7 ms followed by 1 IPI at 95 ms], and the CTX inputs consisted of a 20 Hz Poisson pulse train. Further, the TCR neuron received inhibitory inputs from the reticular nucleus (RN) and thalamic inhibitory interneurons (TIN) represented using biophysical cable models (Fig. S2B). The RN axons were driven by excitatory inputs from the TCR neurons and CTX (Fig. S2B), and the TIN axons received excitatory inputs from both CTX and CER (Fig. S2B). The balance of the synaptic strengths of the CTX and CER inputs to the TCR neuron determined which of three types of firing behavior each TCR cell exhibited (Fig. S2C): (1) burst (oscillating in theta band), (2) regular firing, and (3) random firing. The proportion of tremor (burst) vs. non-tremor (regular and random) TCR cells in the VOP, VIM and VC TH were based on published data [28]: 50% burst, 30% regular, and 20% random (Fig. S2D).

### Spatial distributions of tremor cells in the computational model

The locations of tremor cells within the TH were based on published microelectrode recordings from patients with ET [16]. To assess the impact of the location of the tremor cells on model LFPs and theta power, we quantified model LFPs with three different distributions of tremor cells: (1) normal distribution, (2) clustered distribution and (3) uniform distribution. For the first case, the location of each tremor cell was derived from a normal distribution with mean and standard deviation from published human data [16] : (1) 5.2 ± 2.1 distance anterior to PC, (2) 4.0 ± 2.1 distance inferior/superior to AC-PC line, (3) 13.5 ± 1.1 distance lateral to the third ventricular wall. For the clustered distribution, the location of each tremor cell was based on the exact location provided in the Brodkey et al. study obtained by digitizing data from the paper [16]. For the third case, the tremor cells were uniformly distributed throughout the rectangular prism. The non-tremor cells were uniformly distributed in all three cases.

### Computation of LFP and modeling deep brain stimulation

Extracellular field potentials at each contact of the modeled bipolar pair of electrodes were computed from the transmembrane currents in each compartment (*j*) of each model TCR neuron (*i*), *I*_membranei,j_ (Fig. S3), using the equation,

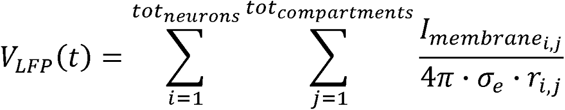

where *r*_i,j_ is the distance of each TCR neural element from the center of the recordingcontact and *σ_e_* is the isotropic and homogeneous conductivity of the extracellular medium (0.3 S/m). We computed the LFP of the bipolar pair, intended to mimic the experimentally recorded LFP, by subtracting the LFPs recorded at the individual positive and negative contacts. The simulated LFP was then processed using the same filter settings as the experimental data (fourth-order Butterworth bandpass filter with a passband of 0.3-500 Hz), and the Welch algorithm was used to compute power spectral density. Fraction of power in the theta band was computed by summing power in the 4-8 Hz band and normalizing by the total power between 1 to 55 Hz. Similarly, the 8-12 Hz band was used to calculate the fraction of power in the alpha band.

DBS was delivered through the modeled electrode located in between the bipolar contact pair used for LFP recordings. The stimulating contact was approximated as a point current source [29]. Extracellular potential *V_e_*(*i*,*j*) due to the point current source at each compartment (*j*) of each model neuron (*i*) was computed using the equation,

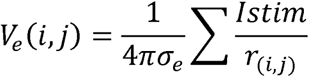

where *Istim* is the stimulation current, *r_i,j_* is the distance from the stimulating electrode to each compartment of the model neuron, and *σ_e_* is the isotropic and homogeneous conductivity of the extracellular medium (0.3 S/m) [30]. All stimuli were biphasic pulses (cathodic first) with a fixed width of 90 µs/phase and an intensity of 2 mA. DBS was delivered at frequencies of 2, 5, 10, 20, 50 and 130 Hz. To minimize the effects of DBS-induced artifacts, the recorded LFP signal was blanked for a period 0.2 ms prior to stimulation to 1.5 ms post stimulation, as in the experiments. Finally, we computed the difference in spectral power in various bands (delta, theta and alpha) between DBS on and DBS off conditions.

Simulations were implemented in NEURON 7.7 with equations solved using the backward Euler method with a time step of 0.025 ms and a total simulation time of 4 s [31]. The first 1 s of the simulation included a transient response and was excluded from analyses. Since the TCR neurons were not connected to each other, we parallelized the model by simulating each TCR neuron and its inputs on a different processor. Following publication, the code required to replicate the model results will be uploaded to ModelDB.

## RESULTS

### Intraoperative LFP and Tremor

We measured simultaneously thalamic LFP and hand acceleration from participants with tremor undergoing VIM DBS implant or pulse generator replacement to quantify neural biomarkers of tremor. We observed significant correlations between LTP and theta band (4-8 Hz, R = 0.455, p = 0.007) and alpha band LFP (8-12 Hz, R = 0.389, p = 0.023) but not the delta (1-4 Hz, R = - 0.271, p = 0.121), beta (13-30 Hz, R = -0.047, p = 0.378), or tremor harmonic (4-Hz band centered at twice tremor frequency, R = 0.156, p = 0.378) bands (Fig. 2A-D; inter- and intra-participant variability see Supplemental Fig. S4). MLR performed on all of the data revealed weak, but significant predictive power of all power bands to LTP (R^2^ = 0.244, p < 10^-3^, Fig. 2E). We then conducted a simple classification of trials with moderate tremor (LTP > 1 m^2^/s^4^) using a GLM. Using all calculated power bands, the AUC was 0.696 and the majority of the contribution came from the theta and alpha bands with little from the delta and beta bands (AUC 0.671, 0.575 and 0.494 respectively; Fig. 2F).

**Figure 2:**
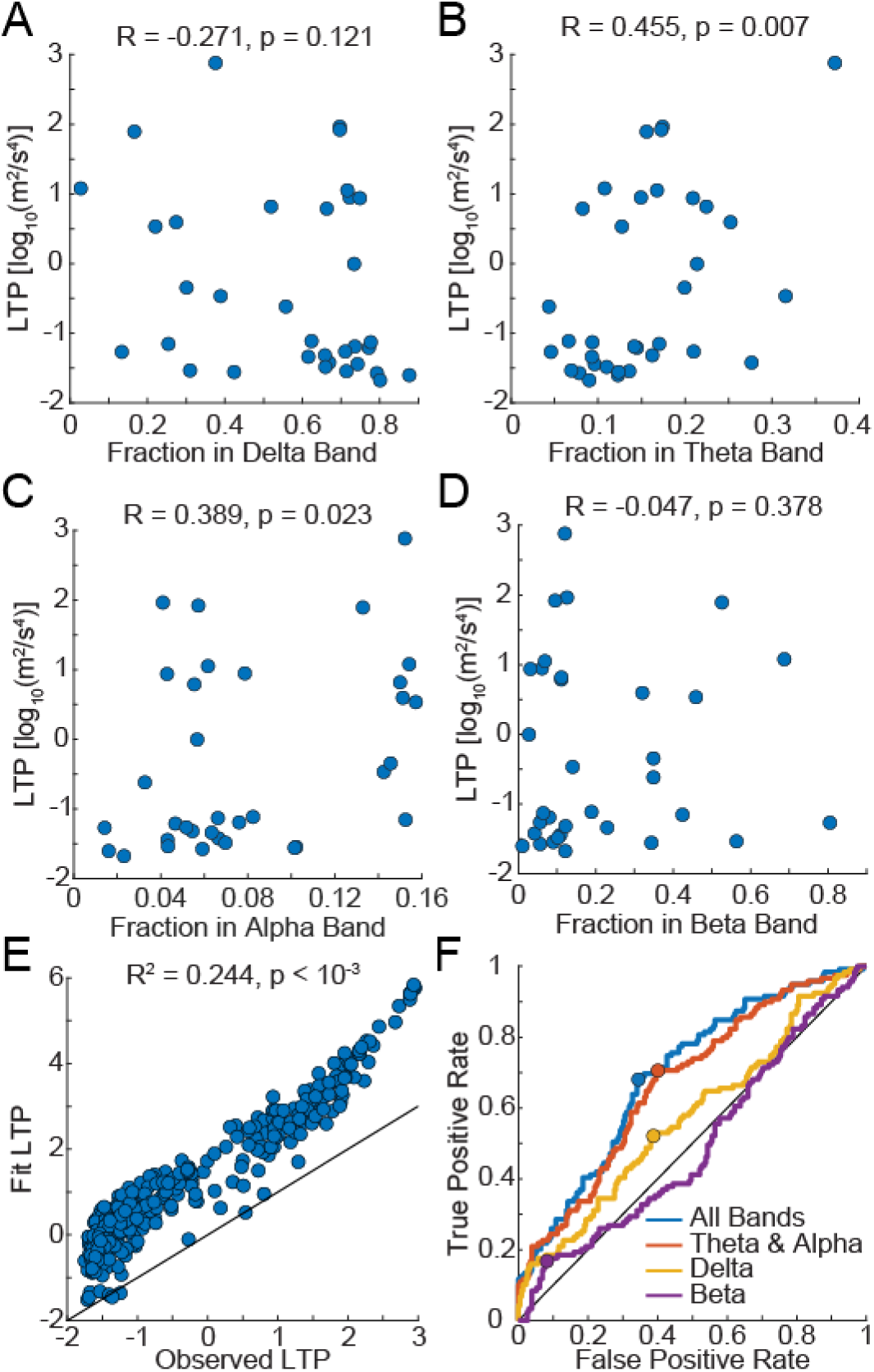
Correlations between LTP and power in LFP in the absence of DBS. The LTP vs the fraction of the total LFP power in the delta (1-4 Hz, A), theta (4-8 Hz, B), alpha (8-12 Hz, C), and beta (13-30 Hz, D) bands. R and p values indicate Spearman correlation. E. Observed vs predicted LTP from a multiple linear regression (MLR) trained on all calculated power bands. The black line indicates 1:1. F. Receiver operator characteristic curves for generalized linear models (GLM) used to classify high (> 1 m^2^/s^4^) and low LTP. Dots indicate the best operating point for each GLM.

We next considered the second-by-second changes in tremor intensity and LFP power using MS coherence. The peak MS coherence values between the thalamic LFP and the tremor signal were typically in the theta to alpha range (Fig. 3A), and the peak was often the same or double the observed peak tremor frequency (Fig. 3B). MS coherence had a high correlation with LTP for those participants with moderate tremor during the intraoperative testing (R = 0.559, p < 10^-3^; Fig. 3C). We observed significant correlation between MS coherence and theta power (R = 0.343, p = 0.004; Fig. 3D), but no correlation between MS coherence and alpha power (R = -0.090, P = 0.455; Fig. 3E). Theta power was moderately related to tremor power when compared across participants (average power, Fig. 2) and within each trial (coherence, Fig. 3). However, even when power in multiple bands was combined, the ability of the LFP to classify tremor remained moderate.

**Figure 3:**
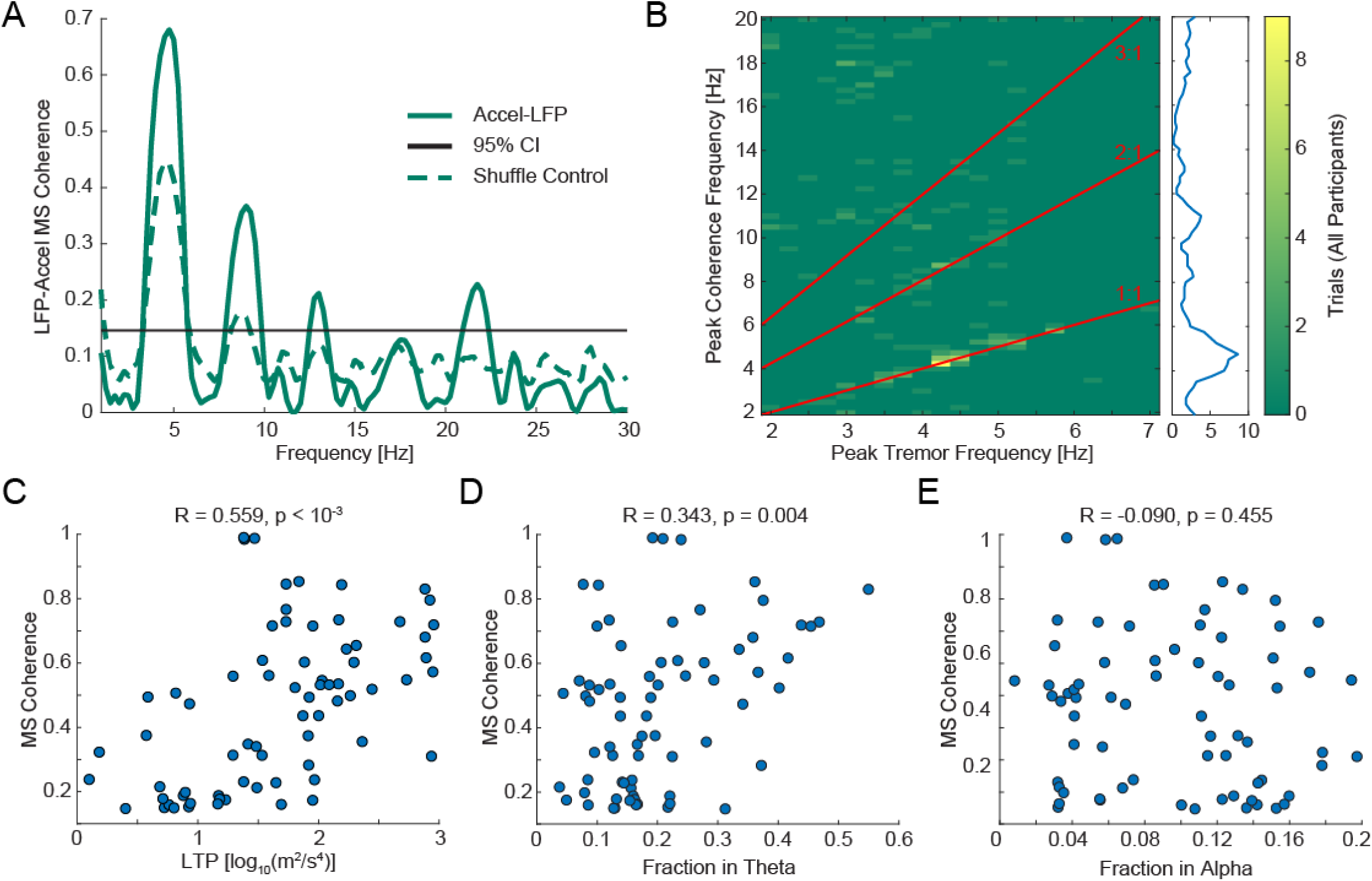
Analysis of coherence between TH LFP and tremor power. A. Example magnitude squared (MS) coherence spectra for a single trial. The black line indicates the 95% confidence interval. Those frequencies exceeding both the confidence interval and the value of the shuffle control were deemed significant. B. The frequency with the greatest significant coherence vs the frequency with the highest amplitude tremor. Values tended to fall on 1:1 and 2:1 ratio lines. Inset: frequency distribution of all maximum coherence values. C-E. The maximum coherence vs LTP, theta band power, and alpha band power for all trials for participants with LTP greater than 1m^2^/s^4^. There was a significant correlation between tremor power and coherence.

DBS was delivered at different frequencies in a subset of participants (n = 16) to determine whether stimulation that modified tremor also resulted in changes in potential LFP biomarkers (Fig. 4A & B). We observed a significant decrease in LTP for 130 Hz DBS across all participants (Fig. 4C, p < 10^-3^). 2, 5, and 20 Hz DBS were observed to increase LTP while 130 Hz decreased LTP in participants with LTP < 1 m^2^/s^4^ (n = 8, one-sided RMANOVA; Fig. S5). Theta band power was increased by 10 and 20 Hz but not by 2, 5, 50 or 130 Hz DBS (Fig. 4D, Table S1). Alpha band power was increased by 2, 5, 10, and 20 Hz DBS and reduced by 130 Hz DBS (Fig. 4E, Table S1). Changes in delta, beta, and tremor harmonic power bands with DBS are contained in the supplement (Supplementary Materials Fig. S6 and Table S1). We tested the correlation between tremor power and each LFP power band for 130 Hz DBS and DBS off. Only theta power was significantly correlated with LTP (Spearman’s R = 0.433, p = 0.014; Fig. S7). We then conducted a MLR to test the predictive power of the fraction of power in bands on the tremor power with 130 Hz DBS. The goodness of fit was moderate but significant (R^2^ = 0.429, p = 0.009) indicating LFP power bands were predictive of tremor, but they explained less than half the variance in tremor power (Fig. 4F). Despite the correlation between MS coherence with LFP without DBS, DBS either increased or decreased the number of trials with significant LFP MS coherence, depending on the frequency (Fig. 4G). We then examined the maximum values (at any frequency of tremor and LFP) of MS coherence during 20, 50 and 130 Hz DBS. 20 Hz DBS often increased low values of MS coherence to well above the significance threshold (Fig. 4G). However, during 130 Hz DBS, no MS coherence greater than 0.3 was observed (Fig. 4H). Together, these data suggest that postural tremor might not be adequately represented in VIM LFP as recorded from DBS leads.

**Figure 4:**
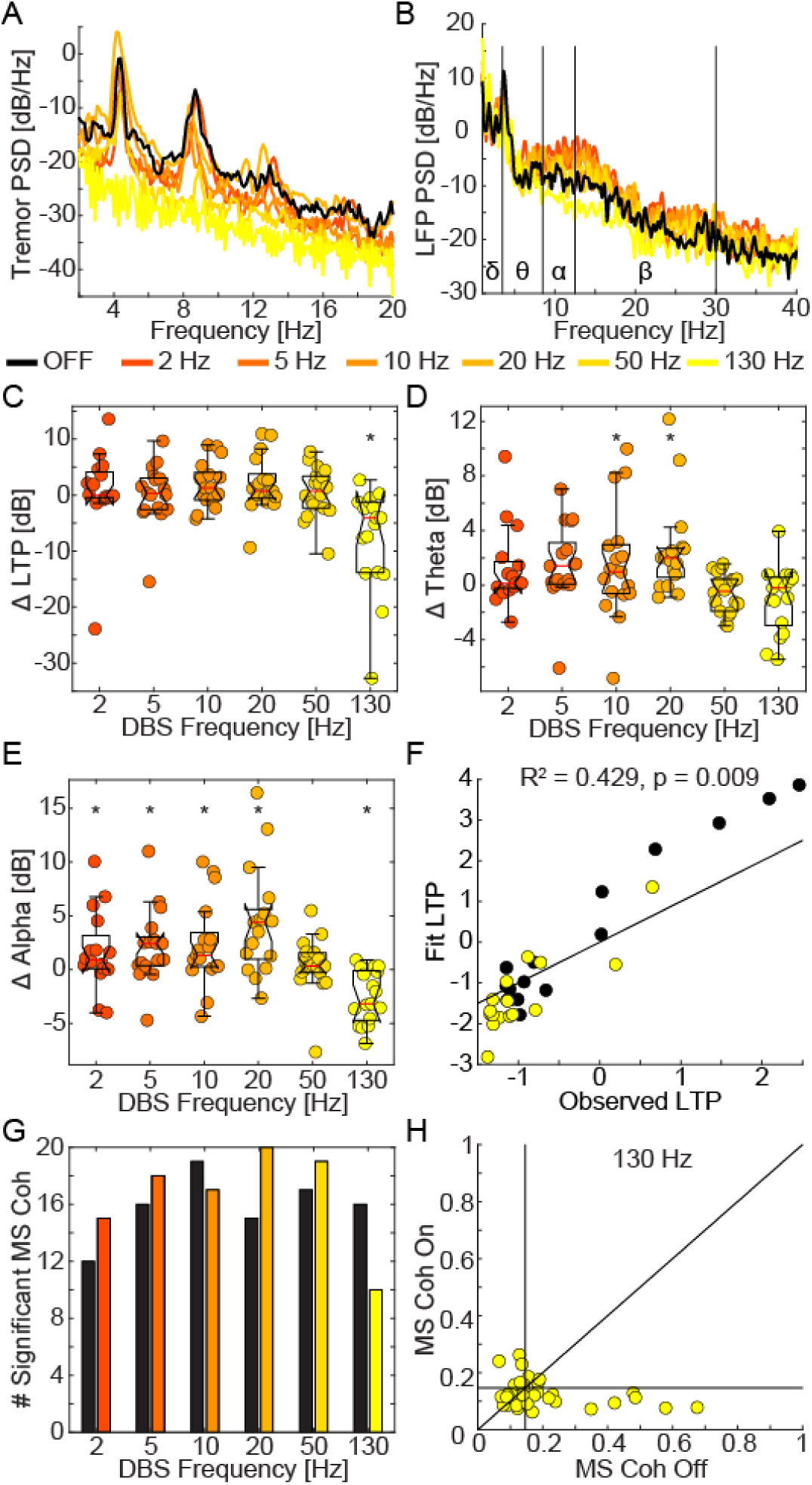
The effects of DBS on tremor and TH LFP. A. Example tremor spectral density taken from the trials of one participant. B. Example LFP power spectral density (same trials as A). C-E. The change in LTP, theta band power, and alpha band power between DBS off and experimental DBS for all participants in the stimulation cohort (n = 16). Boxplot edges are the 25^th^ and 75^th^ percentiles of the data, red lines indicate the means, notches indicate 95% confidence interval of the median, and whiskers cover the range not considered outlier data. F. Fit of a MLR on change in LFP band power versus observed LTP during 130 Hz DBS and DBS off. Black line represents 1:1. G. Total number of trials with significant MS Coherence for DBS off (black) and each frequency of DBS (red to yellow) H. All maximum MS Coherence values during 130 Hz DBS vs maximum MS Coherence values during DBS off. Vertical and horizontal lines denote significant coherence threshold. Diagonal line indicates no change with DBS. * Indicates significant difference from DBS off (RMANOVA, see Methods). MS Coh indicates MS Coherence. R^2^ goodness of fit of the MLR.

### Effect of DBS Electrode Locations on Simulated LFP

We simulated LFP in the computational model and computed spectral power for each bipolar contact pair positioned in the model as in the 34 participants to assess the variation in LFP spectral power with recording electrode location (Fig. 5A1). The tremor cells were distributed normally within the TH with positions derived from microelectrode recordings in humans [16]. The character and spectrum of the simulated LFPs varied based on the location of the DBS electrode relative to the volume of synchronously firing tremor cells (Fig. 5A2, B, C). The fraction of LFP power in the theta and alpha bands varied considerably across the 34 electrode locations (Fig. 5D), and the median fraction of power in the alpha band was lower than the median fraction of power in the theta band (Fig. 5D). Further, the range of theta and alpha in the model agreed well with the range seen in the experimental data (Fig. 2B, C and Fig. 5D1, D2). Finally, we grouped electrodes based on the fraction of power in the theta band to assess any relationship with the location of the recording electrode (Fig. S8). Electrodes exhibiting higher theta power tended to be more superior to the AC-PC line with shorter anterior distance to the PC point than those with lower theta power (Fig. S8).

**Figure 5:**
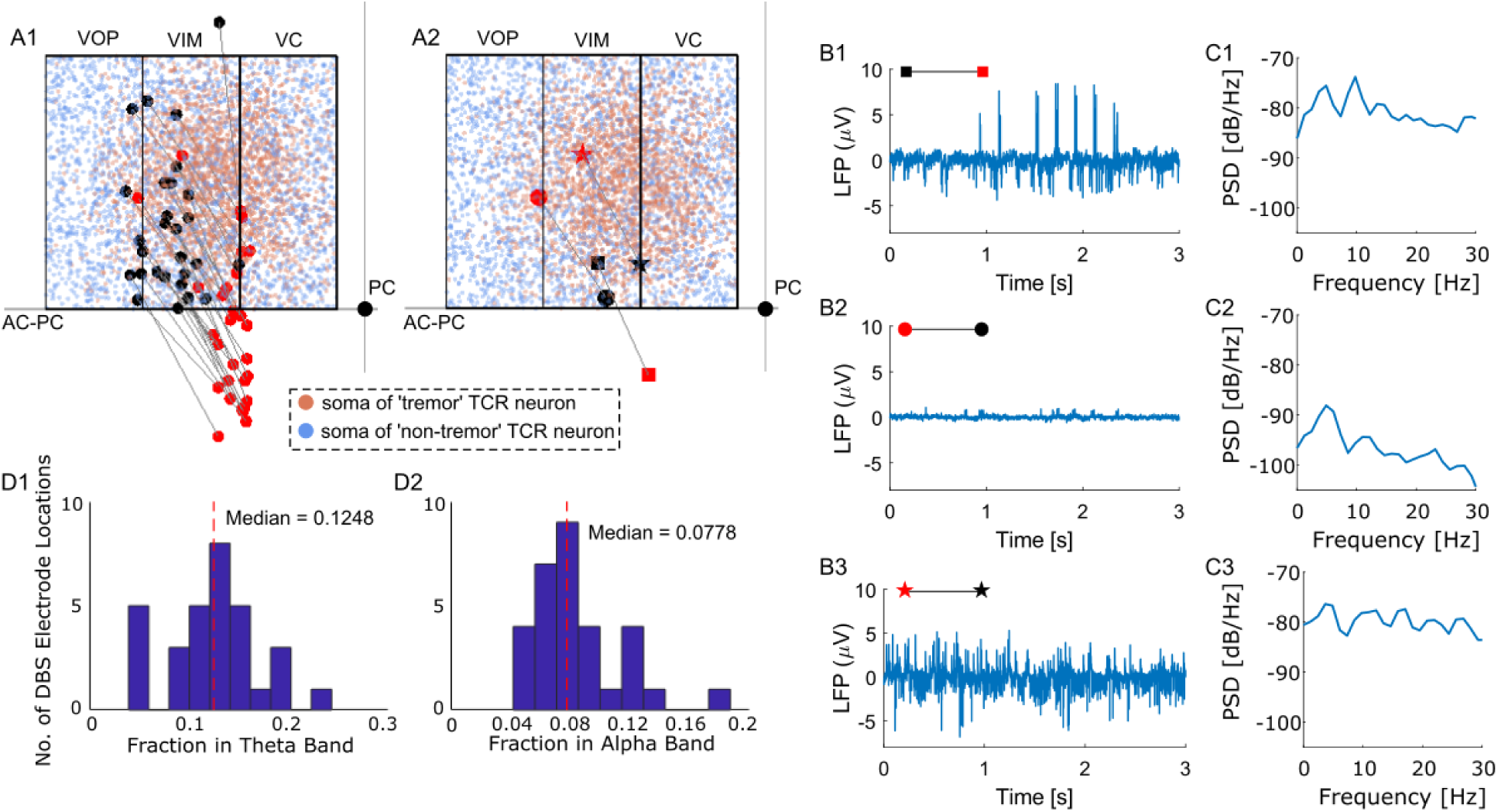
Characteristics of the model TH LFP. (A1) Location of bipolar contact pairs (filled red circle – positive contact and filled black circle – negative contact) in the TH model used to compute LFP. These electrode locations are those from the human LFP recordings. Orange dots indicate the locations of the soma of tremor cells and blue dots represent the locations of non-tremor cells. (A2) Representative examples of three bipolar contact pairs from the human LFP recording locations corresponding to the data in B and C. (B) Simulated LFPs across the three bipolar contact pairs shown in A2. (C) Power spectral density (PSD) of the LFPs shown in B. (D) Fraction of LFP power in theta and alpha bands across all electrode locations shown in A1.

### Effect of Distribution of Tremor Cells on Simulated LFP

We evaluated the impact of three different spatial distributions of tremor cells in the TH on the recorded theta power: (1) normally distributed tremor cells (Fig. 6A), (2) clustered distribution of tremor cells (Fig. 6B), (3) uniformly distributed tremor cells (Fig. 6C). For a given bipolar contact pair (with theta fraction of power >0.15 with the normally distributed tremor cell condition), the power in the theta band varied substantially based on the distribution of the tremor cells in the TH (Fig. 6D).

**Figure 6:**
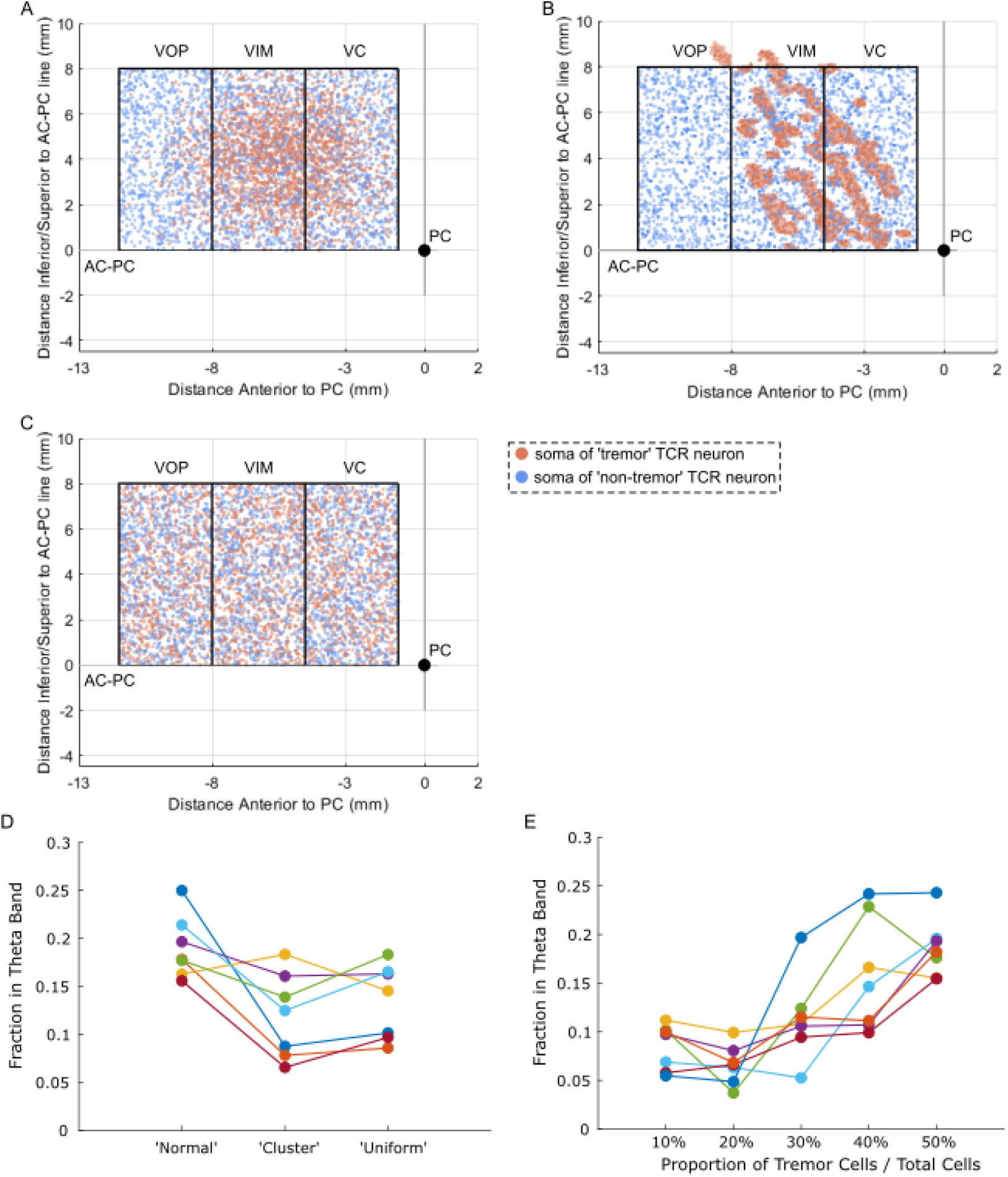
Effect of the spatial distribution of tremor cells on theta power in simulated LFP. The location of tremor cells (shown in orange) was based on (A) normal distribution, (B) clustered distribution, and (C) uniform distribution. (D) Fraction of LFP power in theta band across distribution types for only those bipolar contact pairs (shown in Fig. 5) with fraction power in theta band >0.15 for the normally distributed condition. (E) Fraction of LFP power in theta band for different ratios of tremor to non-tremor cells. Each color trace represents a different contact pair, and only those electrode locations with fraction of power in theta band >0.15 at 50% tremor to non-tremor cell ratio are shown here. The tremor cells were normally distributed. The effect of DBS frequency on theta power for these electrode locations (i.e., fraction theta power >0.15) is shown in Fig. 7.

### Effect of Proportion of Tremor Cells on Simulated LFP

We quantified the theta power in the simulated LFP signal as a function of the proportion of synchronously firing tremor cells by reducing the ratio of tremor cells to non-tremor cells from the original 50%, while the proportion of uniformly distributed non-tremor cells was increased to maintain a total count of 5000 TCR neurons. The fraction power in the theta band decreased with a reduction in the ratio of tremor to non-tremor cells for electrode locations with theta fraction of power >0.15 at the original 50% proportion (Fig. 6E).

### Effect of DBS Frequency on Simulated LFP

We quantified the effects of DBS frequency on simulated LFP spectral power in the delta, theta and alpha bands for a subset of bipolar contact pairs from the human LFP recording locations. For electrode locations with a high theta fraction of power (>0.15) during DBS off, high frequency DBS (130 Hz) decreased power in the delta and theta bands compared to DBS off consistently across electrode locations (Fig. 7 A, B, C, D). In contrast, delta and theta power varied considerably across electrode locations during low frequency DBS (Fig. 7C, D). Further, power in the alpha band varied substantially across electrode locations at all DBS frequencies (Fig. 7E). For electrode locations with low theta fraction of power (<0.15) during the DBS off condition, there was no consistent modulation of LFP spectral power in the delta and alpha bands across electrode locations at any of the tested DBS frequency (Fig. S9). Although 130 Hz DBS decreased theta power, the extent of decrease was comparable to 2 Hz stimulation for some electrode locations (Fig. S9D). These results indicate that the theta fraction of power during DBS varies with electrode location (i.e., across patients), and LFP spectral power may not be a reliable biomarker for tremor for both DBS on and off conditions.

**Figure 7:**
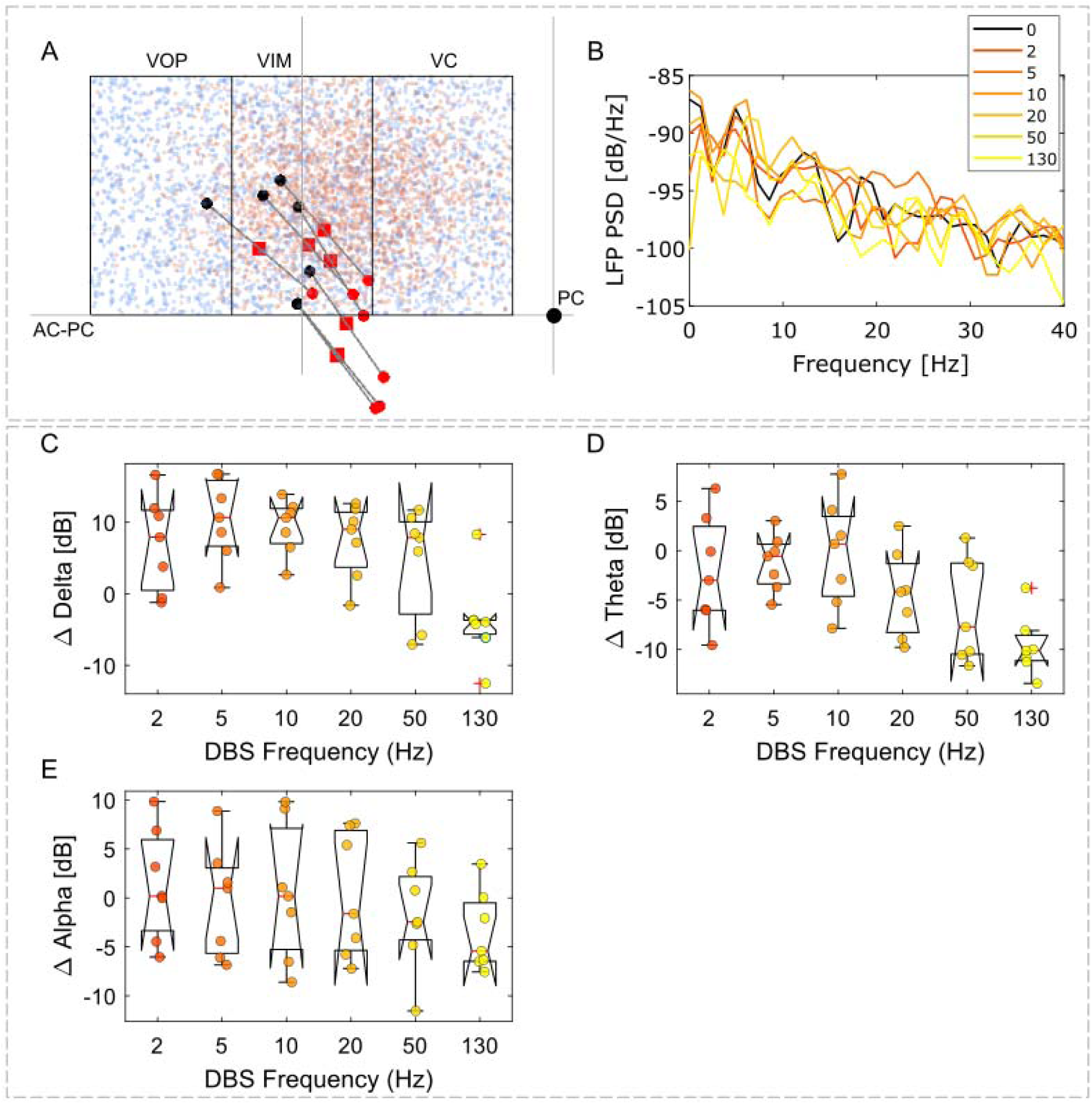
The effects of DBS on simulated thalamic LFP in the model. (A) Location of DBS contact used for stimulation (red squares) and the corresponding bipolar contact pair used for recording LFP. Only those electrode locations with fraction of power in the theta band (>0.15) during the DBS off condition were included. Orange dots represent tremor cells and blue dots show non-tremor cells. (B) Representative simulated LFP PSD across DBS frequencies recorded at one of the electrode locations shown in A. (C-E) The change in delta band power, theta band power, and alpha band power between DBS off and DBS on for the bipolar contact pairs (n=7) shown in A. Boxplot edges are the 25^th^ and 75^th^ percentiles of the data, red lines indicate the means, notches indicate 95% confidence interval of the median, and whiskers cover the range not considered outlier data. Each dot on the box plot represents a different bipolar contact pair used for the LFP recording.

## DISCUSSION

We quantified potential biomarkers of tremor amplitude in VIM LFP across a large cohort of movement disorder patients with upper limb tremor. We observed significant correlations between theta and alpha band recorded LFP power and measured tremor power. However, even when multiple power bands were combined, the predictive power of the LFP remained relatively low. 130 Hz DBS significantly reduced LTP but did not strongly modulate power in any of the LFP bands (e.g., theta oscillations). We used a computational model to demonstrate that the theta signal depends critically on the spatial distribution of the population of tremor cells relative to the placement of the DBS electrode, and this provides a potential explanation for the heterogeneity in the human thalamic LFPs.

### Biomarkers for tremor

Tremor can be directly measured and classified using wearable devices [32–36]. However, wearables are devices additional to the DBS system, the required wireless communication greatly decreases battery life, and therefore, an on-device tremor correlate is desirable for adaptive DBS. Several recent studies examined LFPs as biomarkers for adaptive DBS. Patients with ET predominantly present with action and postural tremor [37, 38]. Action tremor is transient, begins at the start of movement and is very common in ET. Recent publications focused on decoding voluntary movement using LFPs recorded from DBS electrodes in TH and/or subdural electrodes over the motor cortex [10, 20, 22, 39, 40]. These studies observed movement-induced beta desynchronization of motor cortical or VIM LFP (and therefore the likely onset of action tremor), and this effect was sustained up to 24 months post-implantation of electrodes. Since we sought to find biomarkers specific to tremor occurring at either posture or rest as opposed to voluntary movements, we did not decode voluntary movements in this study.

Prior studies attempted to decode postural tremor from TH LFPs by correlating LFP spectral power (especially in the theta band) with tremor signal measured using an accelerometer or EMG [10, 17–21, 39, 41]. Pedrosa and colleagues found a moderate correlation between tremor and LFP spectral power in the theta band in a small patient cohort [19, 41]. Further, they saw a strong correlation between tremor and peak LFP-tremor coherence [21]. Across a large patient cohort, we saw a moderate correlation between tremor and LFP spectral power in the theta/alpha bands, which predicted the significant MS coherence observed here and agrees well with the findings by Pedrosa and colleagues [21]. Tan et al. implemented a machine-learning model to decode tremor using LFP spectral power from different bands [20]. Their GLM decoded movement-or-tremor events with an average detection rate of 80% and spectral power in the theta band contributed the most to tremor decoding [20]. Our findings are consistent with their results, with a 70% detection rate of tremor by the GLM and spectral power in the theta/alpha bands substantially contributing to the tremor detection. A recent study by He et al. decoded tremor-provoking states in the presence of 130 Hz DBS with a detection rate of ∼80%, with spectral power in the theta band contributing the most to tremor detection [22]. We found only moderate correlation between tremor and spectral power for theta band during 130 Hz DBS (and paired DBS off trials) in our patient cohort. Further, we found a moderate correlation between true and observed tremor predicted by training a MLR model using spectral power from different bands during 130 Hz DBS. Although studies, including ours, have shown a moderate correlation between LFP spectral power in the theta/alpha bands and tremor with and without DBS, this effect may wane 12-24 months post-implantation [39].

### Effect of tremor cell distribution and DBS electrode location on LFP

TH LFP appears to be only a modest biomarker for tremor with theta power correlating moderately with tremor. Our modeling results suggest the limited correlation seen in experimental studies could be due to variations in theta power associated with the specific spatial distribution of tremor cells relative to the recording electrodes. The distribution of tremor cells within the TH remains unclear, with some studies suggesting a clustering of tremor cells at the VIM-VC border [16, 19, 42], while others indicate a more widespread distribution throughout VOA, VOP and VIM, with the location of tremor cells not being predictive of DBS electrode location to treat tremor [43]. Further, the theta power in the model varied with DBS electrode location for a given distribution of tremor cells. There is no consensus in the field regarding the optimal DBS electrode placement to treat tremor [4]. VIM of the TH used to be the conventional DBS target to treat tremor, but recent studies suggest the posterior subthalamic area (PSA) to be the more ideal target due to its proximity to the cerebellothalamic tract [44–46]. Activation of the cerebellothalamic tract has been implicated in mediating the therapeutic effects of DBS in treating tremor [27, 47, 48]. The location of DBS electrodes varied considerably across patients in our study, with recording contacts spanning VOP, VIM, and PSA regions (Fig. S1).

### Study limitations

Intraoperative experiments have limitations, including limited time, microlesion effect in the case of new implants, and potential influence of anesthesia on the neural recordings. Together these may have weakened the correlations between LTP and its representation in the LFP. Postural tremor may have been limited due to participants being confined to the surgery table, further weakening the correlation between tremor and LFP. Our results suggest a better representation of tremor amplitude in LFP in patients with LTP ≥ 0 m^2^/s^4^. This study used macroelectrodes with recordings performed from contacts unused for DBS. Our computational data suggests that microelectrode recordings could observe changes in thalamic LFP when located near tremor neurons, whereas the intraoperatively LFP responses were necessarily averaged over up to 6 mm, rather than local recordings. Limited recording time precluded us from training a tremor classifier individualized to each participant as in the Tan et al. study [20]. Our classifier therefore exhibits a lower AUC but may be applied to any patient with upper extremity tremor and VIM LFP. However, as adaptive DBS transitions to clinical practice, we envision semi-automatic selection of individualized classifier parameters will be part of initial DBS programming. We did not analyze the effect of disease state on neural biomarkers in this study due to the large skewness in the sample size of ET (n = 25) vs. PD (n = 6) vs. multiple sclerosis (n = 1) patients. Although different pathophysiological mechanisms might underlie tremor symptoms in ET vs. PD patients [49], studies have shown similar electrophysiological findings inside the thalamus in both diseases [19]. Imaging would lead to a 1-2 mm error in estimating the DBS electrode location [16]. This margin of error would have a limited impact on our conclusion on the effect of electrode location on simulated theta power since the recording contact locations in our patient cohort spanned a broader region (> 2mm). We did not systematically record LFPs before and after cued movements; therefore, we could not decode tremor-provoking voluntary movements in the current study. Building on our study’s findings, deciphering neural biomarkers for tremor with chronic in-home TH LFPs recorded using a bidirectional device such as Medtronic percept would represent an exciting path forward.

## Supporting information

Participant Data Table in Supplementary Materials

## Data Availability

All data necessary to interpret, verify, and extend the research will be anonymized and made publicly available with the final version of this article. Following publication, the code required to replicate the model results will be uploaded to ModelDB.

## Acknowledgments

We acknowledge technical support from the Duke Compute Cluster. The authors thank Robert Gross for assistance in collecting intraoperative data.

## Author Contributions

K.K., S.L.S., B.D.S., C.S.O., D.A.T. and W.M.G. conception and design of research; S.L.S., B.D.S., C.S.O., J.J.P., K.T.M., and D.A.T. performed experimental data collection. K.K. and Y.Z. implemented computational model and ran simulations; S.L.S., K.K. and A.V. analyzed data; K.K., S.L.S. and W.M.G. interpreted results; K.K. and S.L.S. prepared figures and drafted manuscript; K.K., S.L.S., Y.Z., A.V., B.D.S., C.S.O., J.J.P., K.T.M., D.A.T. and W.M.G. edited and revised manuscript; K.K., S.L.S., Y.Z., V., B.D.S., C.S.O., J.J.P., K.T.M., D.A.T. and W.M.G. approved final version of the manuscript.

---

**Figure S1:**
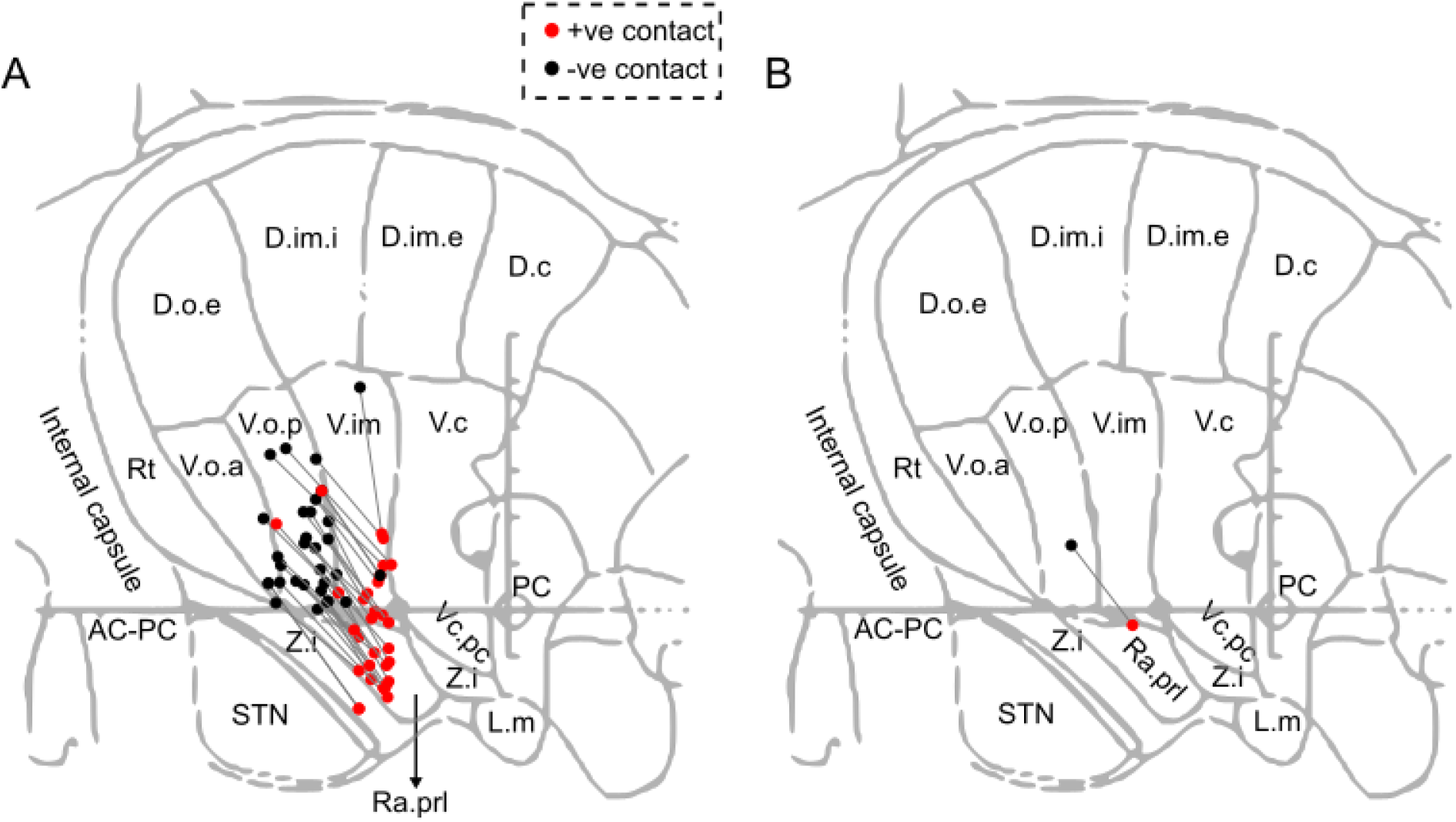
Position of bipolar contact pairs used for recording LFPs across participants. (A) Location of bipolar contact pair across participants. (B) Mean contact location across participants. The mean location of the positive contact across participants was 0.63 mm inferior to anterior commissure (AC) – posterior commissure (PC) line and 4.88 mm anterior to the PC point. Similarly, the mean location of the negative contact was 2.62 mm above the AC-PC line and 7.02 mm anterior to the PC point. The contact location is plotted using distances relative to the PC coordinate and AC-PC line on a digitized sagittal stereotactic map from the Schaltenbrand-Wahren atlas. The anterior distance of contacts was scaled to the standard AC-PC length of 24 mm to account for the differences in intercommissural distance across patients. The trajectory of the DBS electrode was such that the distal contacts were predominately located in the posterior subthalamic area and the proximal contacts on the border of VOP/VIM and in VOP. Activation of the cerebello-thalamic tract is speculated to mediate the therapeutic effects of thalamic DBS for tremor, and the tract runs through the posterior subthalamic area (Gallay et al. 2008).

**Figure S2:**
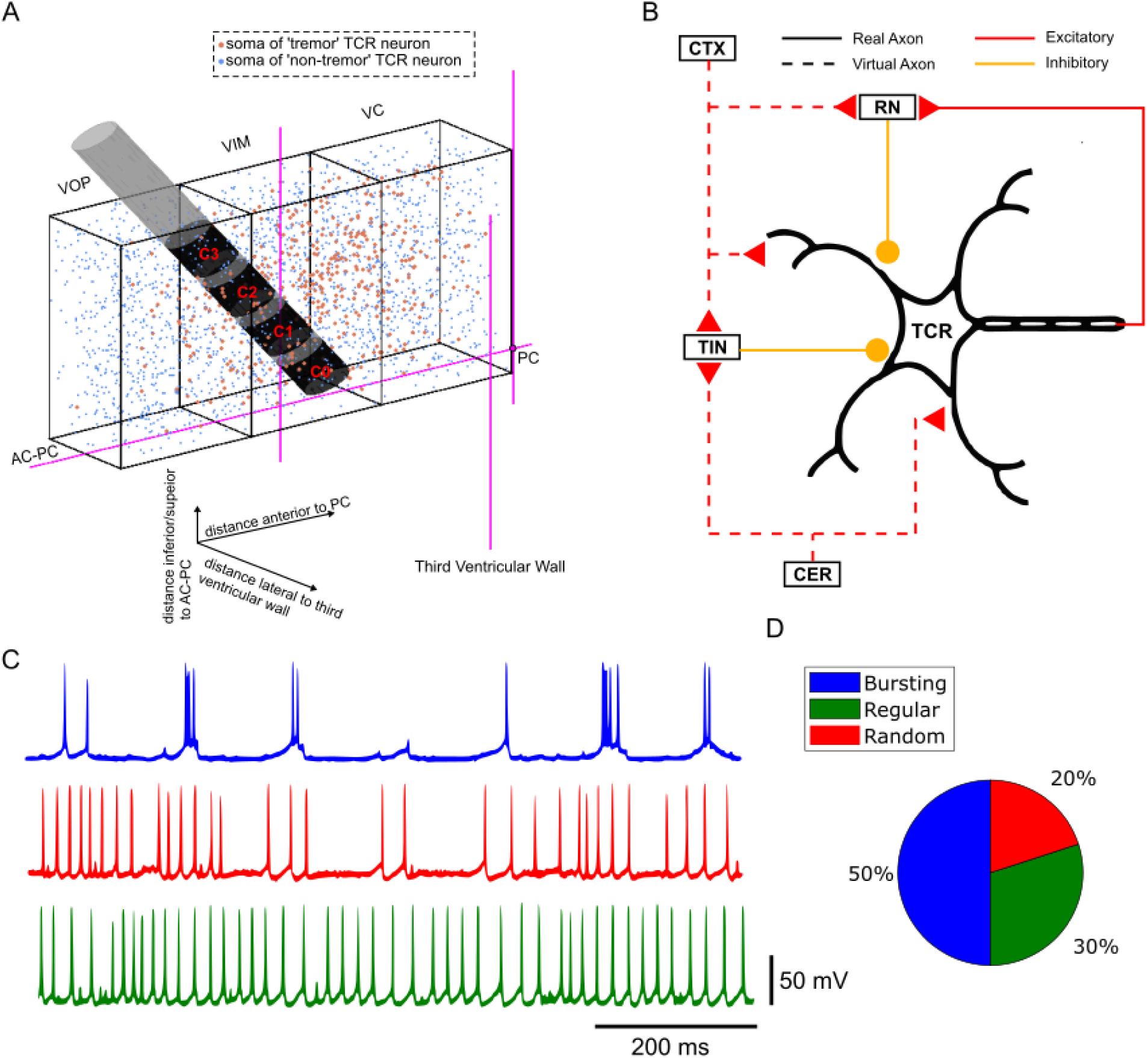
Computational model of thalamic LFP recording. (A) Rectangular prism representation of VOP, VIM and VC regions of the TH showing somata of tremor (orange dots) and non-tremor (blue dots) model TCR neurons. The dimensions of each rectangular prism were 3.5 mm x 4.5 mm x 8 mm. The 3389 DBS electrode was inserted in the VIM region. The tremor cells were normally distributed with the mean and standard deviation determined from microelectrode recordings of tremor cells in humans (Brodkey et al. 2004). The non-tremor cells were uniformly distributed throughout the TH. (B) Schematic of microcircuit of TCR neuron with inputs received from other regions. The TCR model neuron was adapted from a published study (Birdno et al. 2012). The TCR neuron comprised a cell body, 251 dendritic compartments, and a double-cable myelinated axon of diameter 5.7 µm. The TCR neurons received inputs from the cortex (CTX), cerebellum (CER), thalamic interneuron (TIN), and reticular nucleus (RN). Inputs from the CTX and CER were represented using virtual axons. Thalamic interneuron and reticular nucleus inputs were represented via double-cable myelinated axons of diameter 2 µm. There were no connections between the TCR neurons in the model. (C) Firing behavior of tremor and non-tremor model TCR neurons. Tremor neurons fired in burst mode at ∼5 Hz. Non-tremor neurons exhibited either a regular or a random firing pattern. Both regular and random firing neurons were classified as non-tremor neurons. (D) The proportion of tremor and non-tremor neurons (same in all regions of the model) was determined from microelectrode recordings from tremor and non-tremor neurons in ET patients (Molnar et al. 2005).

**Figure S3:**
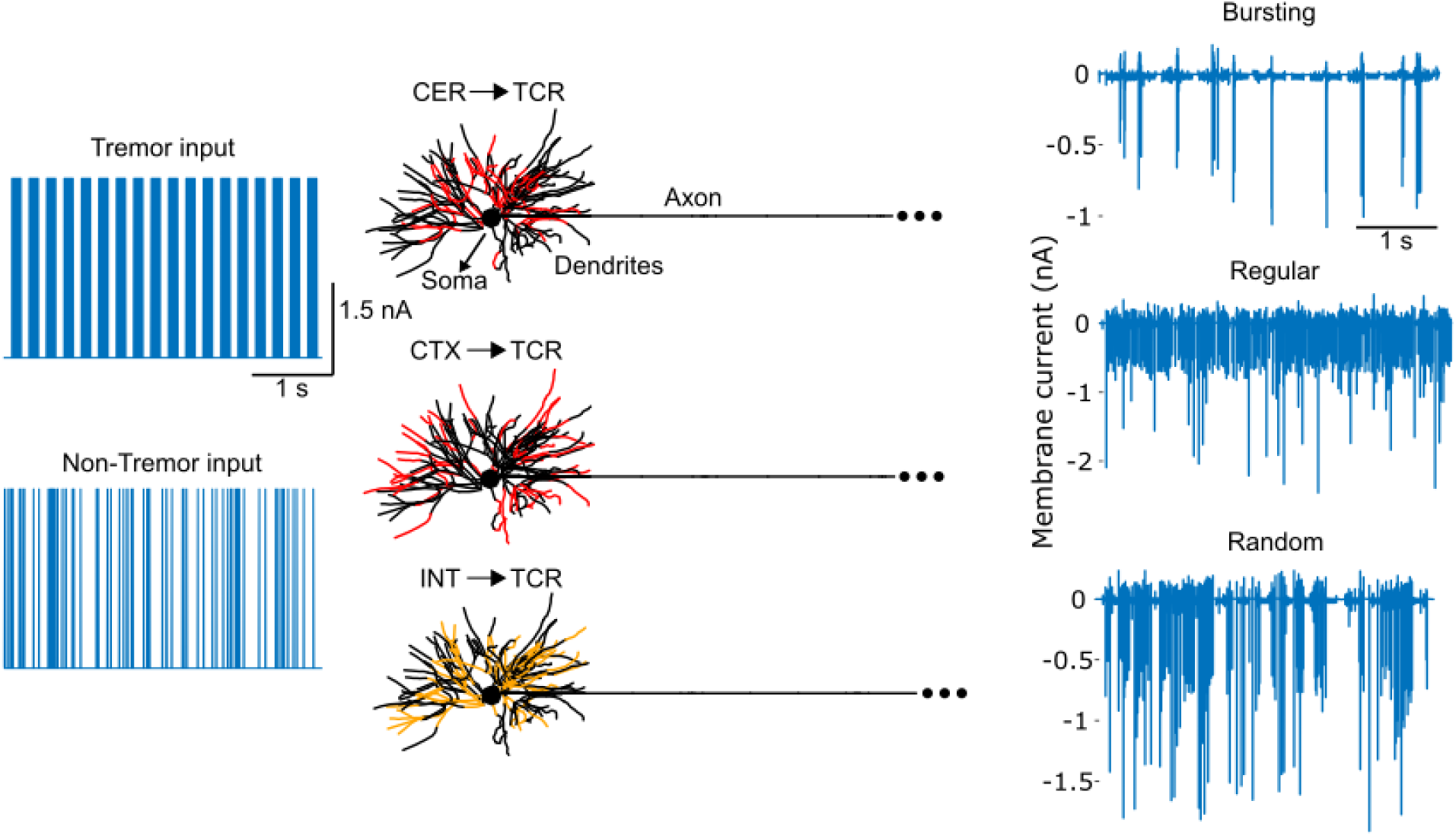
Transmembrane currents in model neurons were the sources that produced simulated LFP in the model. Schematic of multicompartment TCR neuron showing dendrites, soma, and axon. Red dendritic compartments represent the location of excitatory synaptic inputs received by TCR neurons from CER and CTX. Yellow dendritic compartments represent the inhibitory synaptic inputs from TIN and RN. Periodic tremor/burst inputs were fed into CER→TCR synapses and random Poisson trains were fed into the CTX→TCR synapses. Panels on the right show the transmembrane currents recorded from the soma of each of the three TCR neuron types characterized based on the firing behavior: (1) bursting, (2) regular, and (3) random. Although the transmembrane current was recorded across each of the 490 compartments of the TCR neuron, and all of these currents served as the source to compute the LFP at different electrode locations, we show here only the transmembrane current from the somatic compartment.

**Figure S4:**
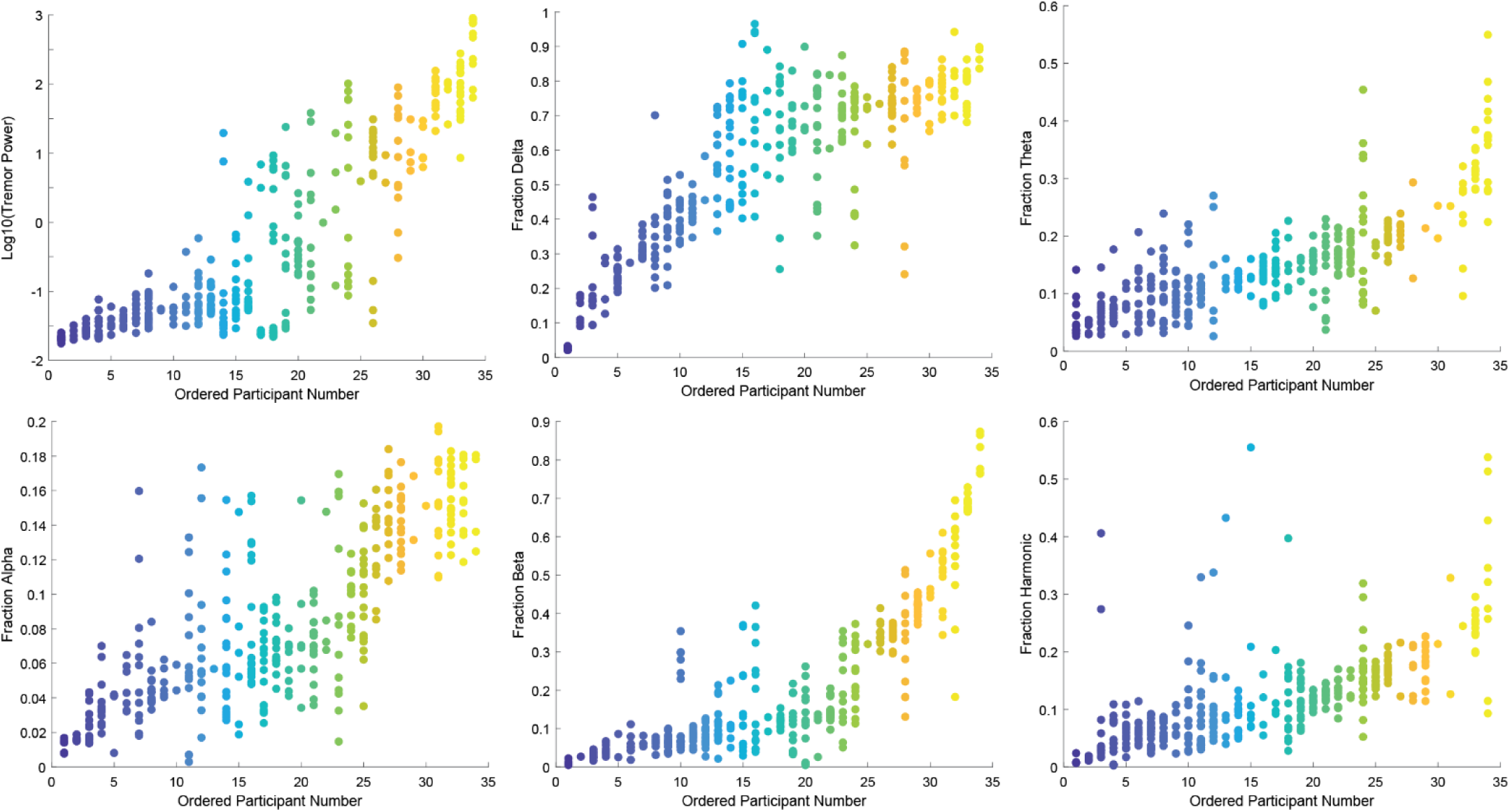
Inter- and intra-participant variability. LTP, delta power, theta power, alpha power, beta power and harmonic power are sorted by participant mean. Each dot indicates a single trial. LFP power bands are normalized by the total power between 2-30 Hz.

**Figure S5:**
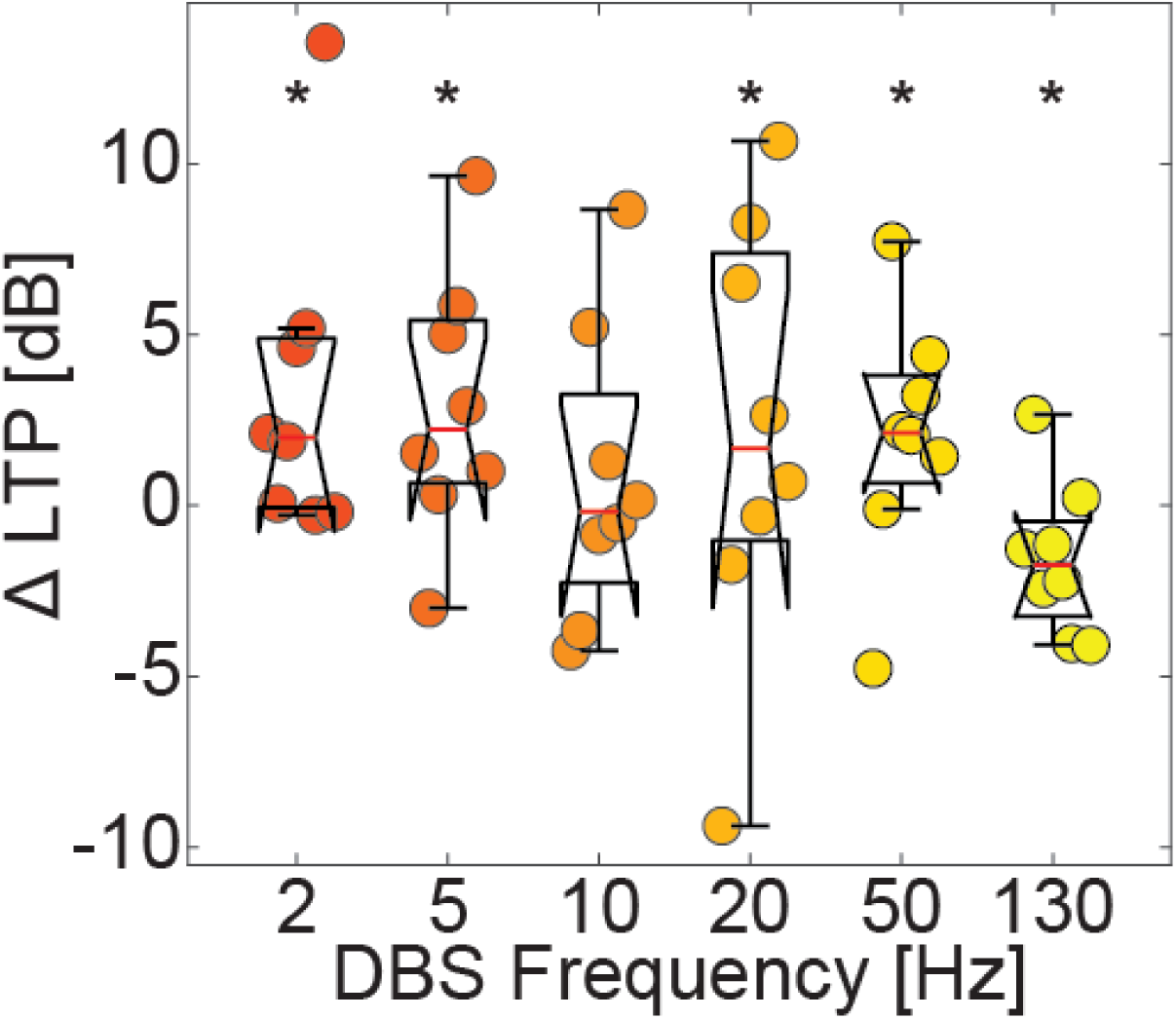
The effect of DBS on LTP in low LTP participants. The change in LTP between DBS and DBS OFF. * Indicates p-values <0.05 calculated by one-sided RMANOVA with *a priori* hypothesis that 2-50 Hz would increase LTP, and 130 Hz would decrease LTP compared to DBS off.

**Figure S6:**
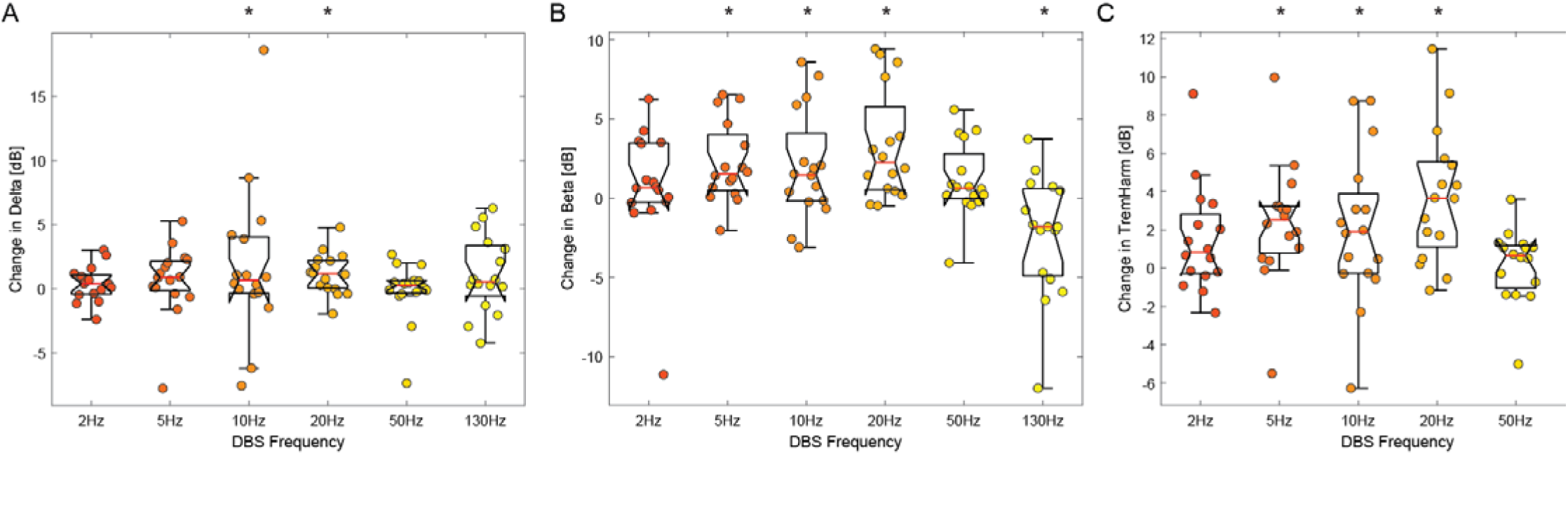
Effect of DBS on delta, beta and tremor harmonic power. A) Change in delta (1-4 Hz) between DBS and DBS off. B) Change in beta (13-30 Hz) power. C) Change in LFP power in a 4 Hz band centered around twice the tremor frequency. * Significant difference from DBS off (2-sided RMANOVA).

**Figure S7:**
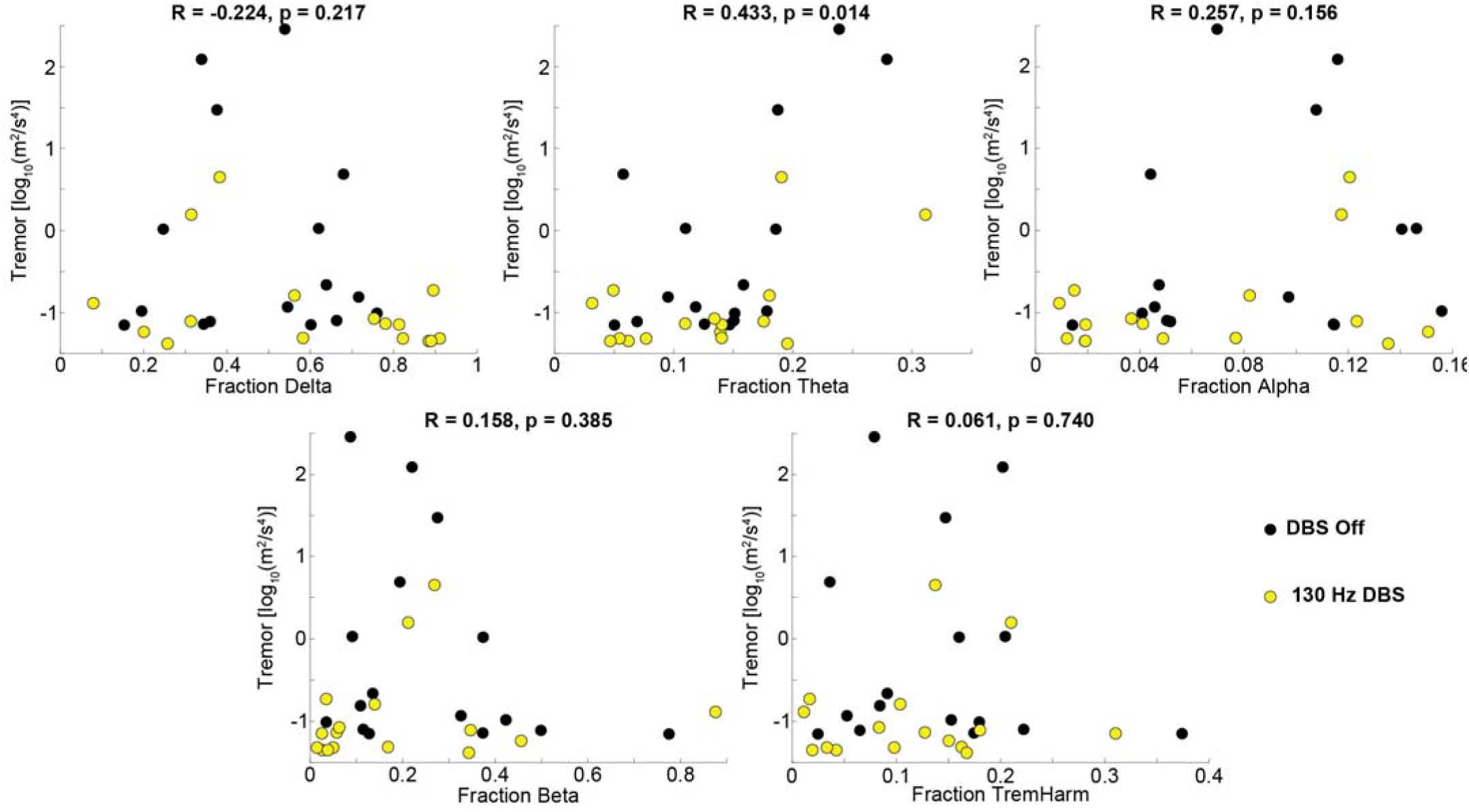
Tremor versus LFP power in frequency bands. The fraction of broadband power in each frequency band for both DBS off (black) and 130 Hz DBS (yellow). Only the fraction in theta was significantly correlated with tremor. Reported R and p values were calculated with Spearman correlation.

**Figure S8:**
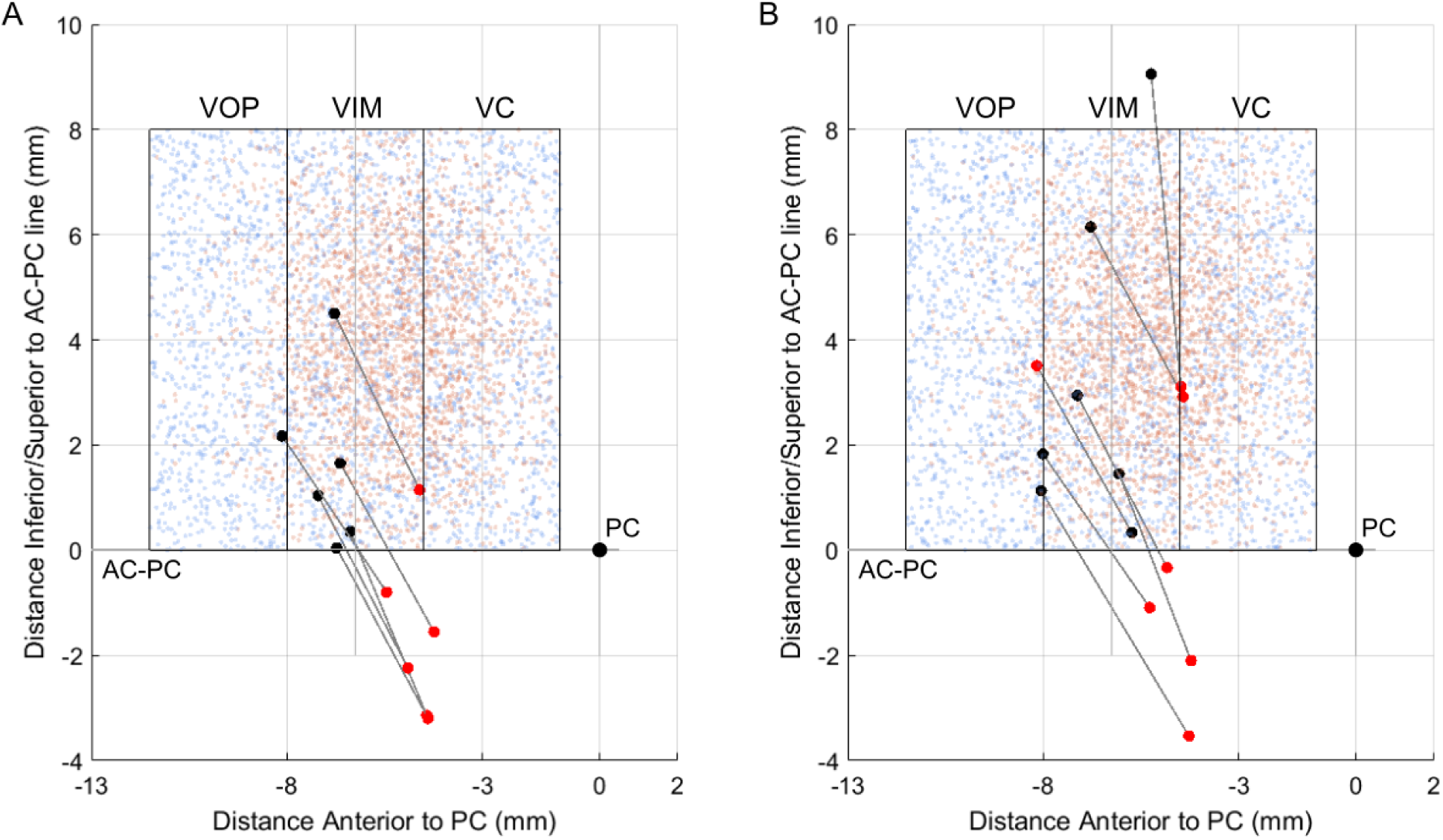
Electrode locations with (A) low fraction power in theta band (<0.085) and (B) high fraction power in theta band (>0.15). Orange dots represent tremor cells and blue dots show non-tremor cells.

**Figure S9:**
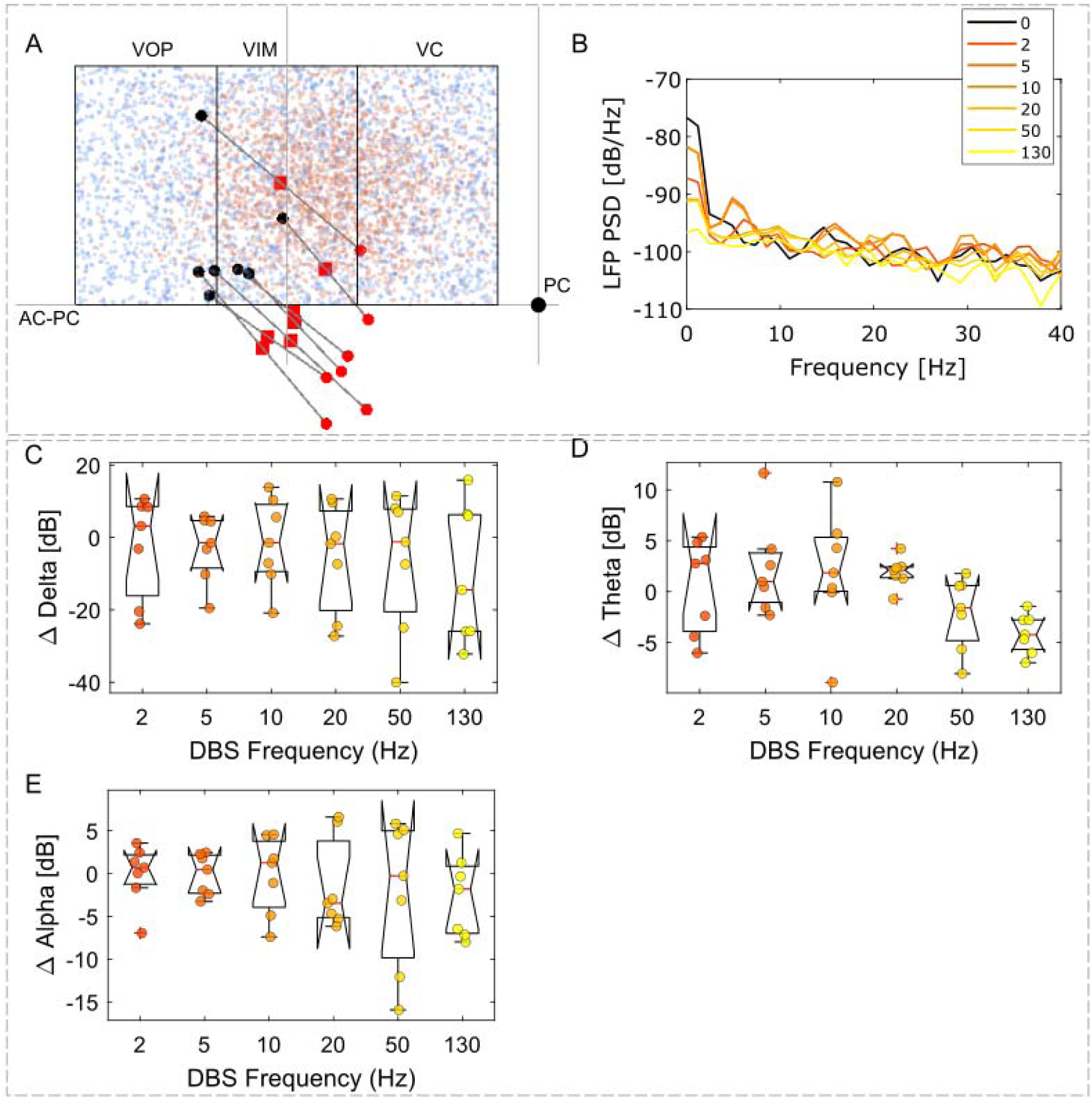
The effects of DBS on simulated thalamic LFP in the model. (A) Location of DBS contact (red squares) and the corresponding bipolar contact pair used for recording LFP. Only those electrode locations with low fraction power in the theta band (<0.15) at the DBS off condition were included. Orange dots represent tremor cells and blue dots show non-tremor cells. (B) Representative simulated LFP power spectral density across DBS frequencies recorded at one of the electrode locations shown in A. (C-E) The change in delta band power, theta band power, alpha band power between DBS off and DBS on for the bipolar contact pairs (n=7) shown in A. Boxplot edges are the 25^th^ and 75^th^ percentiles of the data, red lines indicate the means, notches indicate 95% confidence interval of the median, and whiskers cover the range not considered outlier data. Each dot on the box plot represents a different bipolar contact pair used for the LFP recording.

**Table S1.** RMANOVA p-values (comparison with OFF)

|  | LTP | Delta | Theta | Alpha | Beta | Tremor Harmonic |
| --- | --- | --- | --- | --- | --- | --- |
| 2 Hz | 0.1942 | 0.9938 | 0.1600 | 0.0366 | 0.4849 | 0.2083 |
| 5 Hz | 0.3855 | 0.6359 | 0.0696 | 0.0064 | 0.0625 | 0.0555 |
| 10 Hz | 0.4324 | 0.7720 | 0.0166 | 0.0012 | 0.0206 | 0.0012 |
| 20 Hz | 0.5424 | 0.9910 | 0.0003 | $< 10^{-4}$ | $< 10^{-4}$ | $< 10^{-4}$ |
| 50 Hz | 0.6699 | 0.7318 | 0.9904 | 0.5667 | 0.3173 | 0.7609 |
| 130 Hz | $< 10^{-4}$ | 0.1654 | 0.1840 | 0.0039 | 0.0007 | 0.1761 |

